# Investigations on using Evidence-Based GraphRag Pipeline using LLM Tailored for USMLE Style Questions

**DOI:** 10.1101/2025.05.03.25325604

**Authors:** Tharun Sekar, Kushal, Supprethaa Shankar, Sabah Mohammed, Jinan Fiaidhi

**Affiliations:** Computer Science, Lakehead University, Thunder Bay, Canada; Dept. of Computer Science Lakehead University, Thunder Bay, Canada

**Keywords:** GraphRAG, Neo4j, Pinecone, Retrieval- Augmented Generation, Knowledge Graphs, Question Answering, LLMs, Medical Informatics

## Abstract

The integration of evidence-based reasoning with retrieval-augmented generation (GraphRAG) holds great promise for enhancing large language model (LLM) question-answering (QA) capabilities. This research proposes a GraphRAG framework that improves the interpretability and reliability of LLM-generated answers in the medical domain. Our approach constructs a knowledge graph using Neo4j to represent UMLS medical entities and relationships, and complements it with a vector store of textbook embeddings for dense passage retrieval. The system is designed to combine symbolic reasoning and semantic search to produce more context-aware and evidence-grounded responses. As a proof of concept, we evaluate our system on United States Medical Licensing Examination (USMLE)-style questions, which require clinical reasoning across multiple domains. While overall answer accuracy remains comparable to that of an LLM-only baseline, our system consistently outperforms in citation fidelity — providing richer, more traceable justifications by explicitly linking answers to graph paths and textbook passages. These findings suggest that even when correctness may vary, graph-informed retrieval improves transparency and auditability, which are critical for high-stakes domains like medicine. Our results motivate further refinement of hybrid GraphRAG systems to enhance both factual accuracy and clinical trustworthiness in QA applications.

## I. Introduction

The evolution of artificial intelligence (AI) has significantly impacted information retrieval and question-answering (QA) systems, particularly with the advent of large language models (LLMs). Early research demonstrated the potential of LLMs for medical education, including their performance on standardized exams such as the USMLE. However, initial studies[1] revealed critical limitations, such as factual inconsistencies, hallucinations, and the inability to establish structured reasoning over complex medical knowledge. To address these shortcomings, research progressed toward integrating structured medical ontologies, such as UMLS, into LLM workflows [2], enhancing factual consistency but still facing challenges in dynamic knowledge retrieval.

Recent advancements in Retrieval-Augmented Generation (RAG) have introduced hybrid approaches that combine vector-based retrieval with structured knowledge representations, including knowledge graphs. Studies such as Medical Graph RAG [3] and KRAGEN [4] demonstrate the effectiveness of GraphRAG frameworks in improving contextual grounding and explainability in medical QA. These approaches leverage graph-based reasoning to interconnect medical entities, allowing for more reliable and interpretable responses. However, existing methods still struggle with integrating realworld clinical knowledge, ranking retrieved information effectively, and balancing symbolic reasoning with dense vector retrieval.

Despite advancements in GraphRAG, medical question-answering systems for USMLE-style queries still face key challenges. Current retrieval models primarily rely on either vector embeddings or structured graphs in isolation, leading to issues in relevance ranking, factual accuracy, and contextual coherence. Additionally, existing approaches lack an adaptive mechanism for dynamically weighing knowledge from both sources, often resulting in suboptimal responses that either overfit structured knowledge or misinterpret unstructured clinical text.

This research proposes a hybrid GraphRAG framework designed to enhance medical QA by integrating Neo4j-based UMLS knowledge graphs with a vector store-driven retrieval system. By introducing a graph-enhanced query expansion step and an adaptive re-ranking mechanism, the proposed approach aims to improve answer relevance, factual accuracy, and explainability in high-stakes medical examinations such as the USMLE. The study contributes to the field by bridging the gap between structured and unstructured retrieval, ensuring a more balanced, contextually rich, and clinically aligned QA system.

**TABLE I:**
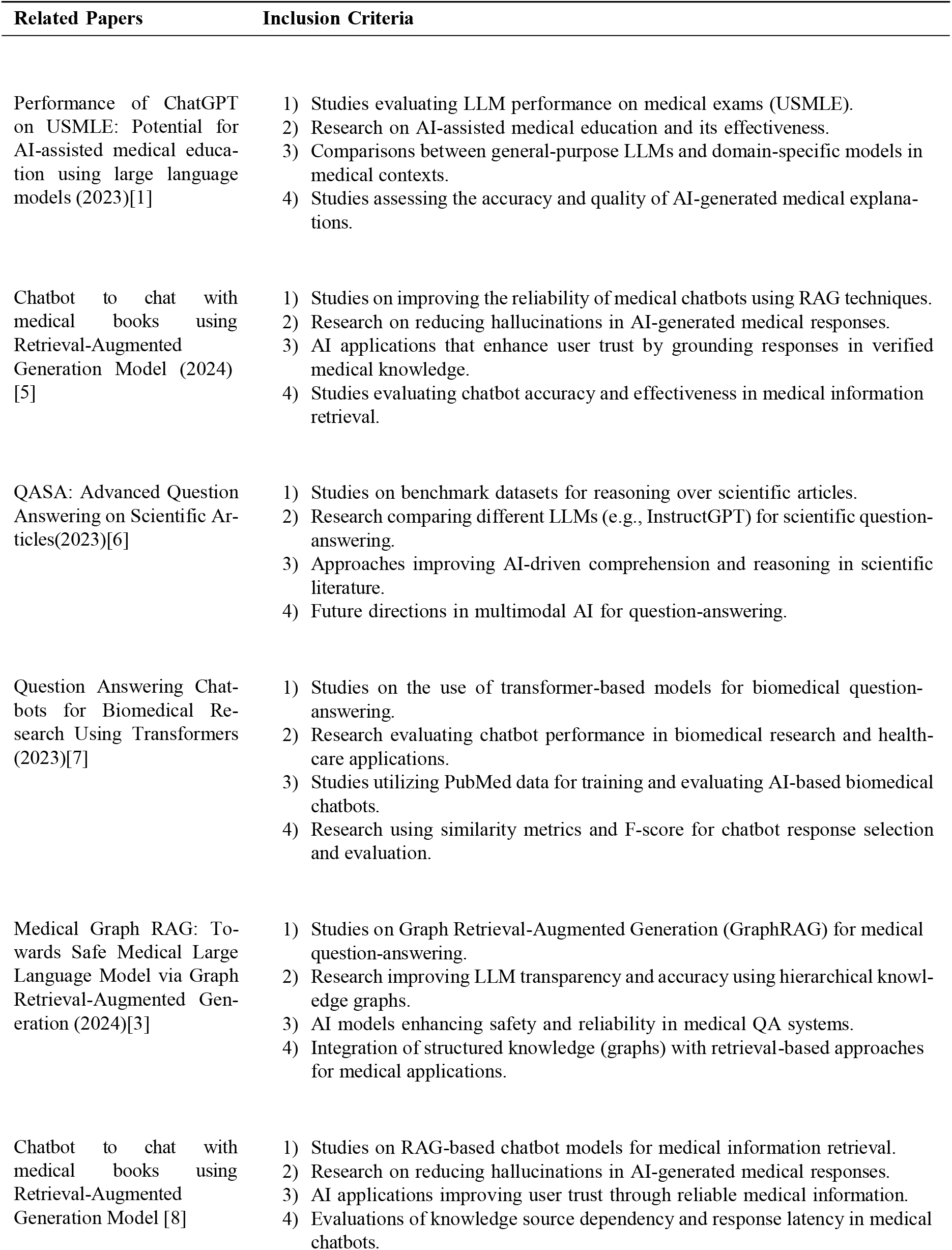
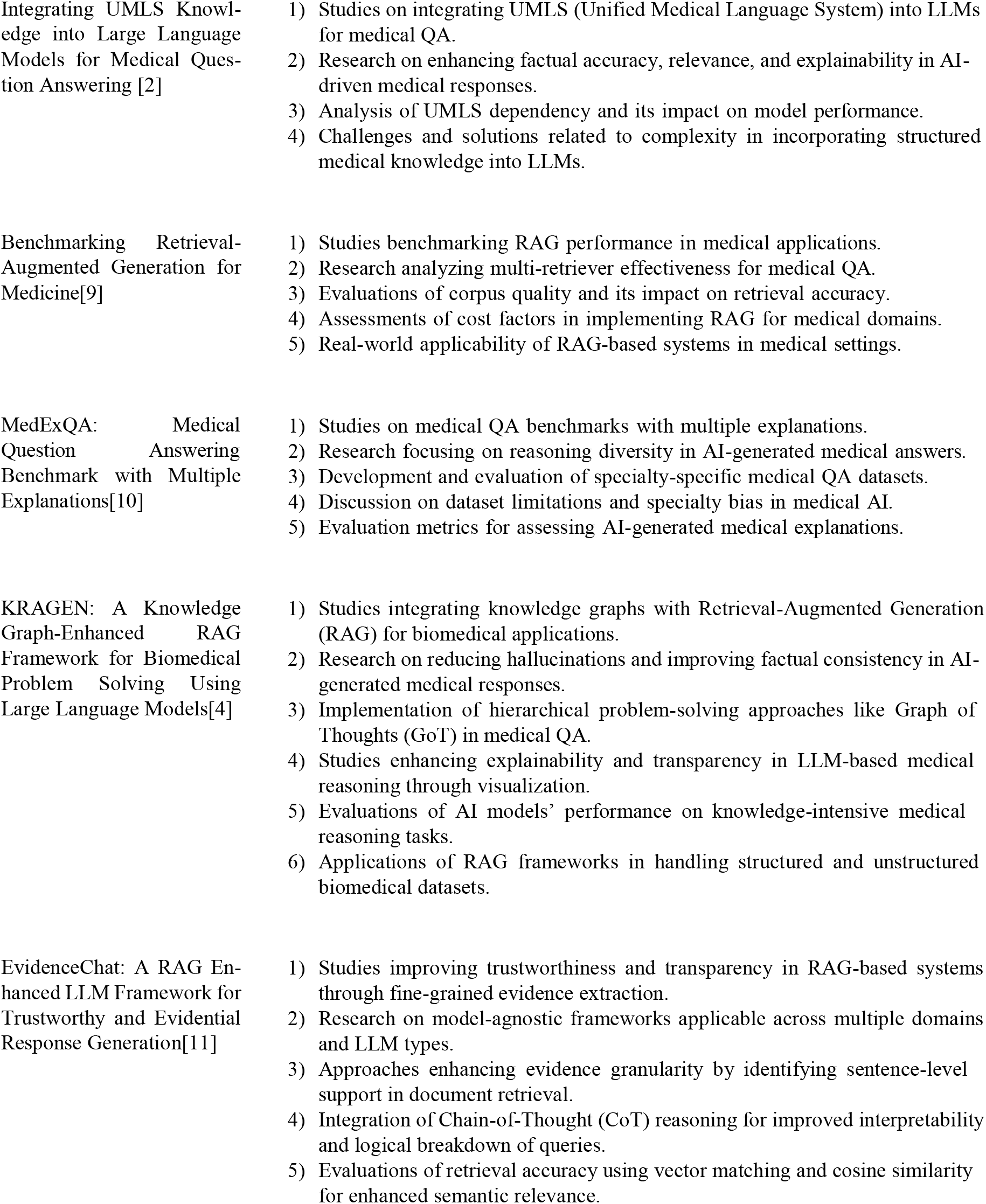
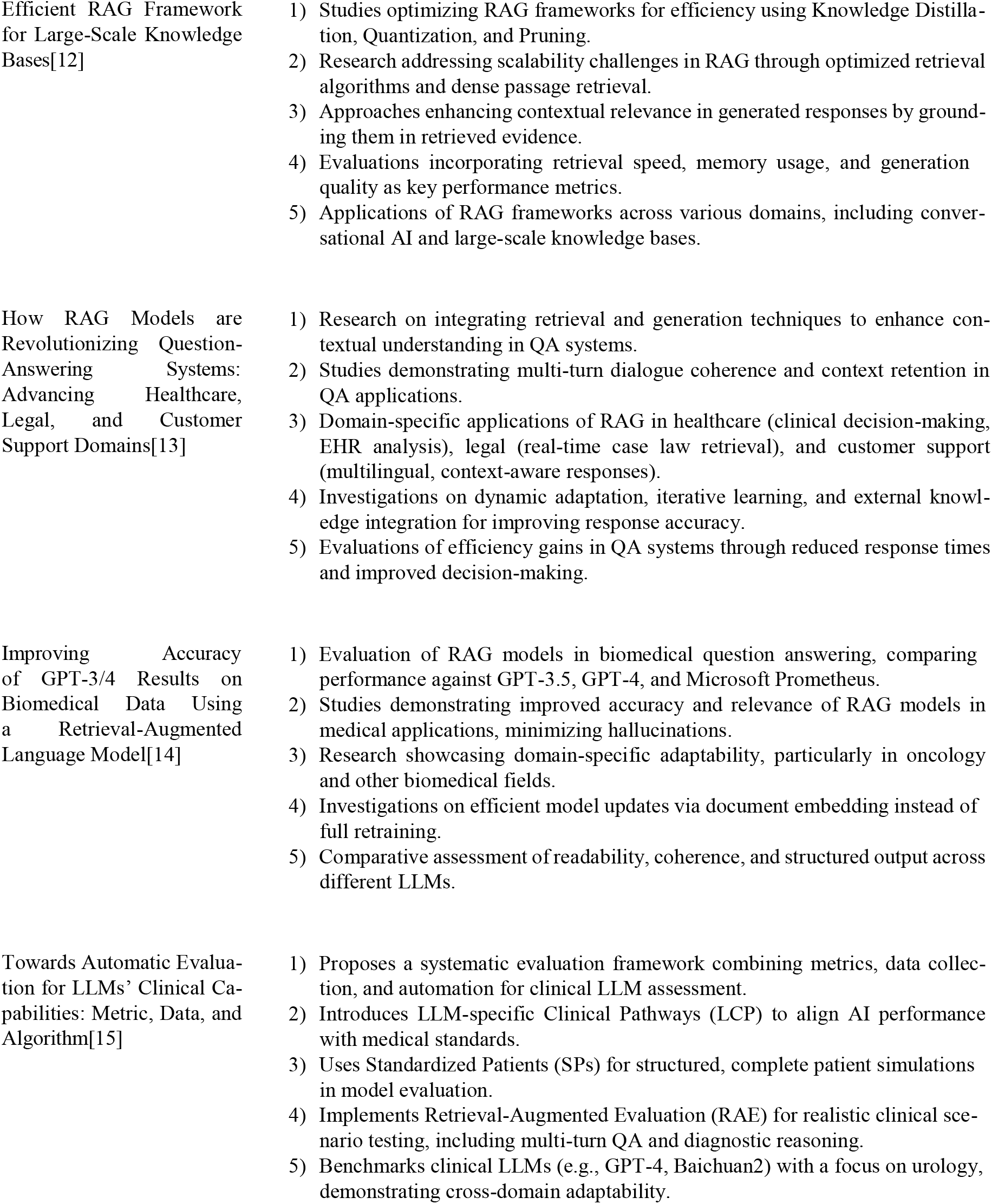
Literature Review Summary.

## II. Literature Review

Early research into the application of large language models (LLMs) in the medical domain demonstrated the potential of AI-assisted education and question answering, particularly for standardized exams such as the USMLE. For instance, Kung et al. [1] evaluated ChatGPT’s performance and highlighted its near-passing capabilities without domain-specific fine-tuning. However, these initial studies also exposed critical limitations, including factual inconsistencies, hallucinations, and a lack of explainability. To address these concerns, later works such as Zhou et al. [2] explored the integration of structured medical ontologies like UMLS into LLM pipelines, improving factual grounding and relevance in medical responses. Despite their contributions, these approaches remained limited by their reliance on either symbolic knowledge or vector-based retrieval alone, lacking a unified retrieval framework to support complex medical reasoning.

More recent and advanced research has begun to explore hybrid architectures that combine both structured and unstructured information sources through Graph Retrieval-Augmented Generation (GraphRAG). Notably, the Medical Graph RAG framework [3] proposes leveraging hierarchical medical knowledge graphs alongside vector-based text retrieval to enhance the safety and transparency of LLM-generated answers. Similarly, the KRAGEN framework [4] introduces a graph-enhanced RAG pipeline for biomedical problem-solving, utilizing multi-hop reasoning over knowledge graphs combined with dense embedding retrieval to improve factual consistency. Additional studies such as EvidenceChat [5] emphasize the importance of fine-grained, sentence-level evidence selection and re-ranking, contributing toward greater trust and interpretability. While these systems advance the state of medical QA, many still face challenges related to retrieval latency, context fusion, and effective ranking of hybrid evidence sources.

The present work builds directly upon these advancements by proposing a practical and balanced GraphRAG pipeline tailored for medical question answering, specifically targeting USMLE-style queries. A comprehensive mind map of the proposed idea is provided in Figure 1. By integrating a Neo4j-based UMLS knowledge graph with a MedQA vector store, this system enables simultaneous symbolic reasoning and semantic retrieval. Unlike prior studies, this pipeline introduces a graph-enhanced query expansion step and an adaptive re-ranking mechanism that weights context from both sources before passing it to the LLM. This approach not only improves factual accuracy and reduces hallucinations but also addresses key shortcomings of previous systems, such as weak interlinking of graph-based concepts and unstructured text. The motivation for this research lies in the growing demand for reliable, interpretable, and domain-aligned LLM applications in high-stakes fields such as medicine, where the synergy of structured and unstructured retrieval can significantly enhance the quality of AI-driven decisions.

**Fig. 1:**
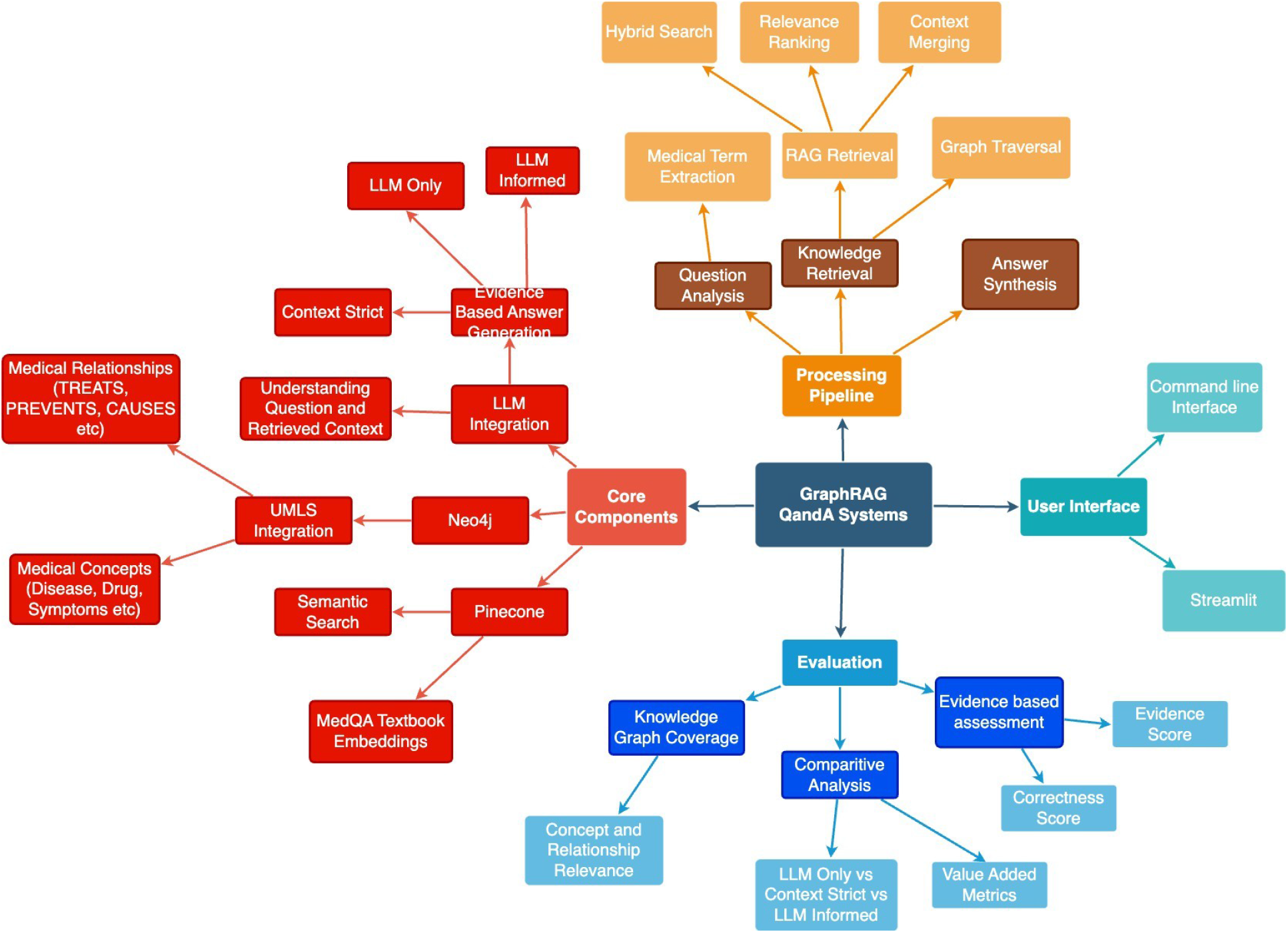
Mindmap of proposed idea

## III. System Architecture and Data Preparation

### A. System Overview

This hybrid GraphRAG system orchestrates retrieval, ranking, and response synthesis through a modular and reproducible workflow. The complete system pipeline (Figure 2) consists of six core stages (Algorithm 1), integrating both symbolic and unstructured data sources for evidence-based medical question answering[16][17]:

**Fig. 2:**
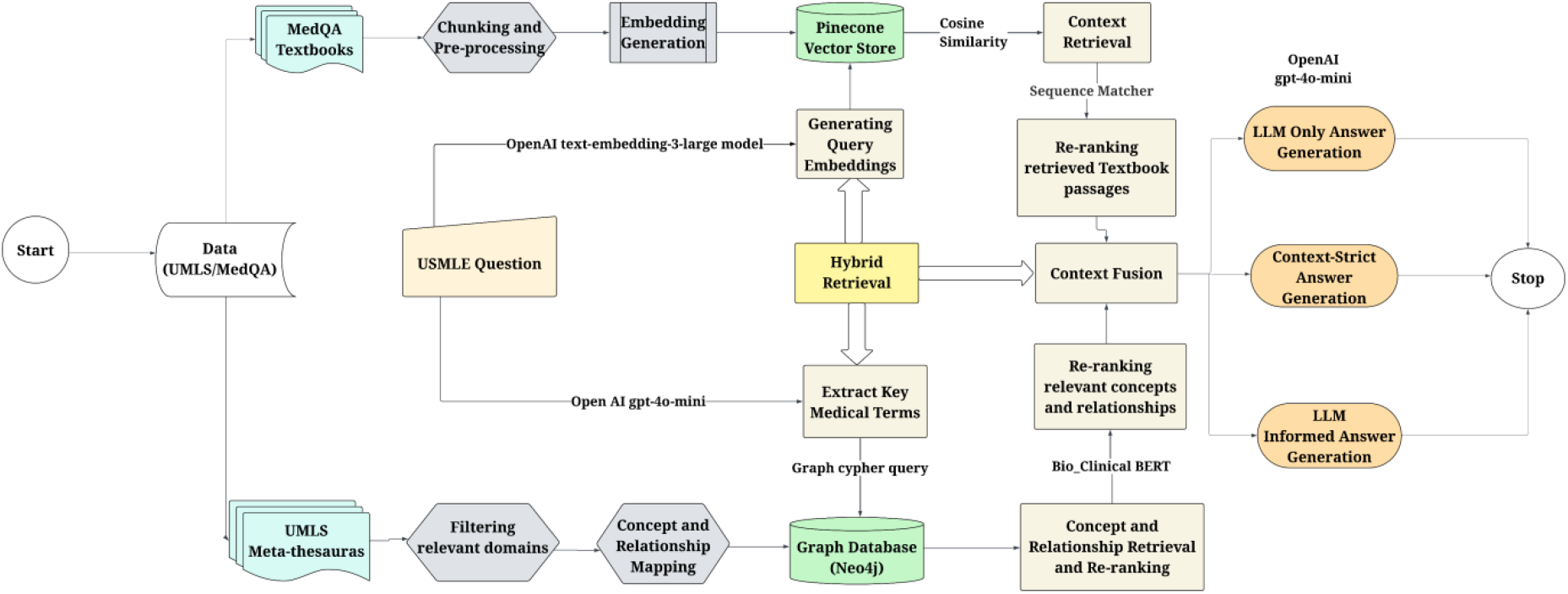
Complete enhanced pipeline diagram illustrating the integration of structured UMLS knowledge graphs and unstructured textbook embeddings within the hybrid GraphRAG framework for evidence-based medical question answering.

**Fig. 3:**
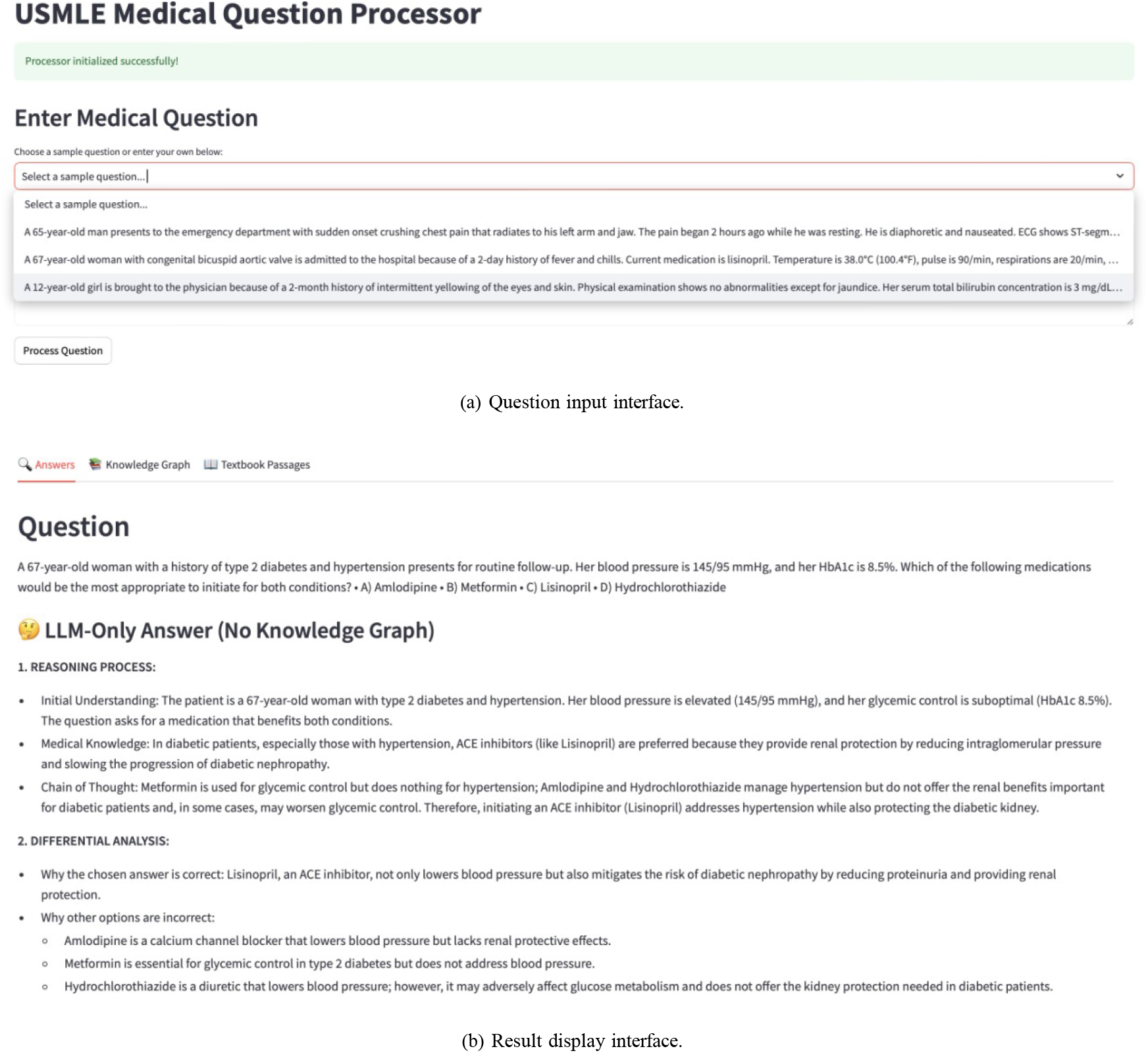
Streamlit-based frontend for medical question processing and evidence-grounded answer generation.

#### 1) User Query Input

A USMLE-style question is entered through a Streamlit-based interface.

#### 2) Query Embedding

The question is embedded using a SentenceTransformer model for vector similarity retrieval.

#### 3) Graph-Based Retrieval

Relevant UMLS concepts and relationships are fetched from the Neo4j knowledge graph.

#### 4) Vector-Based Retrieval

Top-*k* semantically similar textbook sentences are retrieved from Pinecone.

#### 5) Re-ranking and Context Fusion

Both sources are reranked, cleaned, and merged into a final prompt.

#### 6) Response Generation

The prompt is passed to GPT-4o-mini under one of three prompting strategies (LLM-only, context-strict, LLM-informed).

Figure 3 shows the User Interface designed using Streamlit.

##### Algorithm 1

Hybrid GraphRAG Medical QA Pipeline

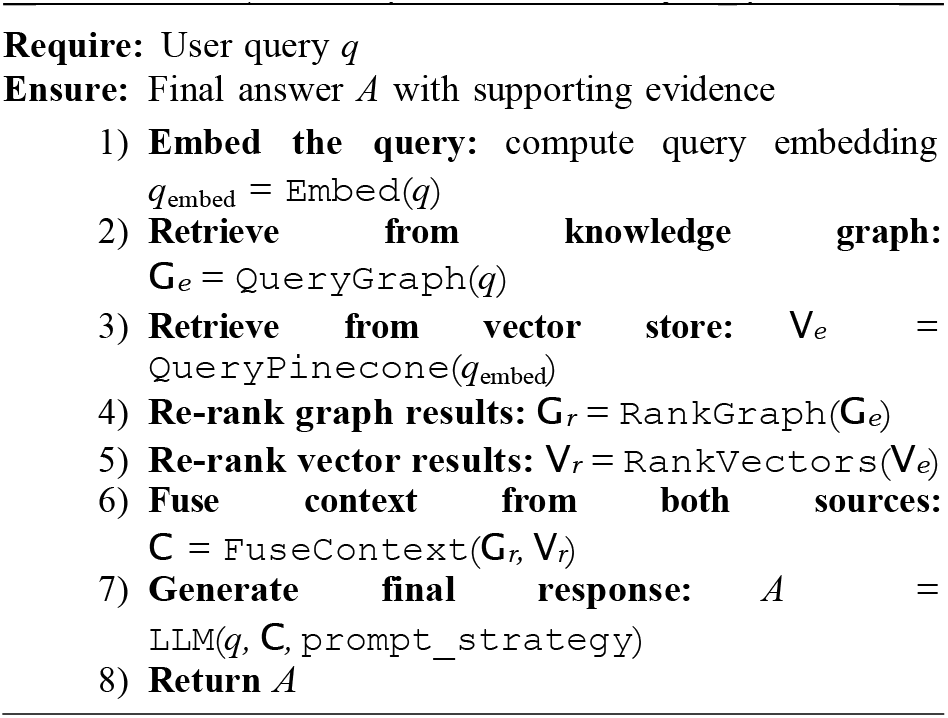

### B. Data Acquisition and Preprocessing

#### 1) UMLS DATASET

The Unified Medical Language System (UMLS) provides a comprehensive source of medical concepts and relationships [18][19]. Figure 4 explains the data acquisition and pre-processing steps in detail.

##### a) Data Cleaning

Preprocessing ensures data integrity through the following steps:

- **Duplicate Removal:** Identified and removed duplicate entries, ensuring each concept and relationship is unique.
- **Inconsistency Resolution:** Addressed conflicting data by cross-referencing authoritative medical sources.
- **Missing Data Handling:** Inferred missing data points from related concepts or used default values to maintain completeness.

**Fig. 4:**
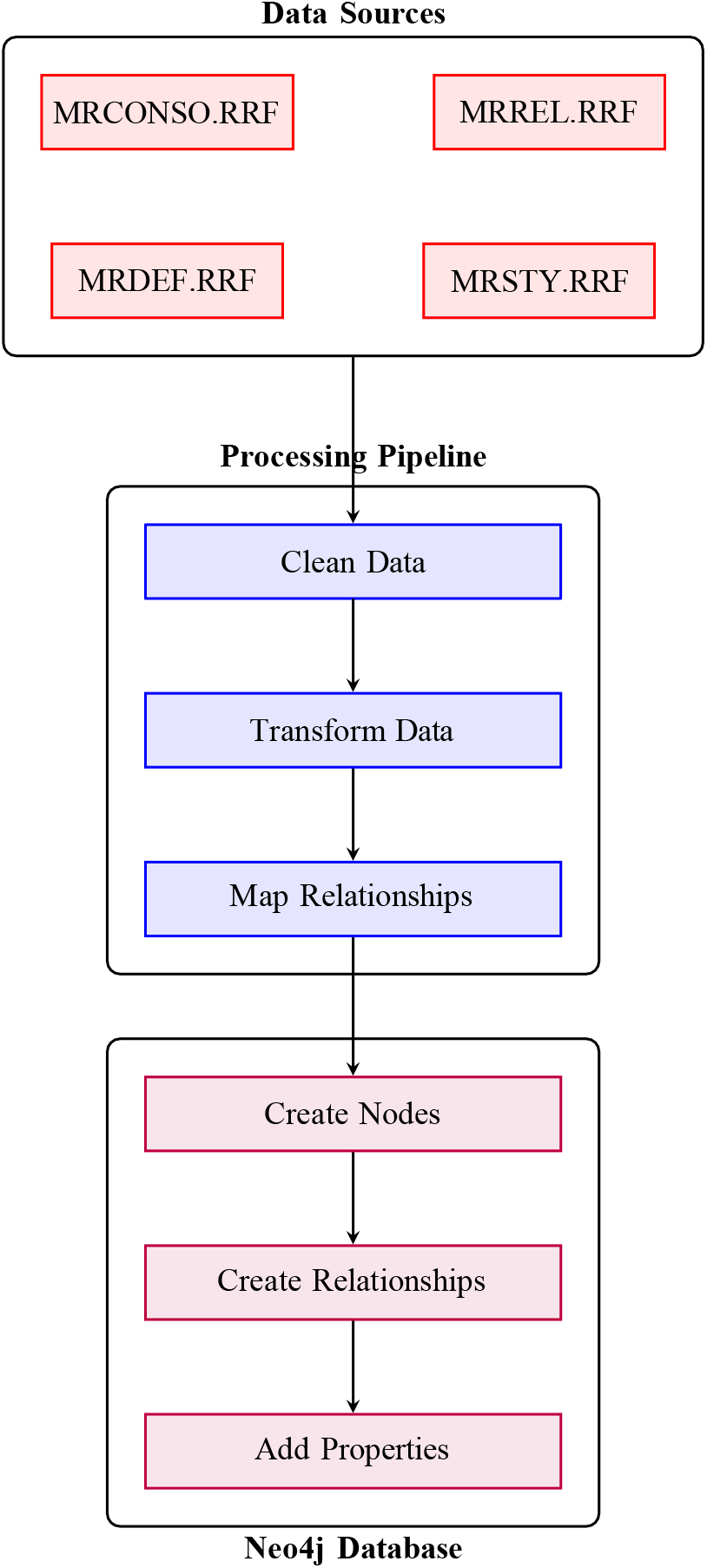
Schematic representation of the graph construction process, showing integration of UMLS datasets into Neo4j.

##### b) Data Transformation

- **Concept Mapping:** Mapped UMLS concepts to graph nodes using their CUIs, assigning labels based on semantic types.
- **Relationship Mapping:** Transformed UMLS relationships into graph relationships using `RELATION_TYPE_MAPPING`.
- **Definition Integration:** Used MRDEF to enrich nodes with definitions, providing contextual information.
- **Property Enrichment:** Added properties such as `term`, `definition`, and `semantic_types` for enhanced concept representation.

#### 2) MedQA DATASET

The MedQA dataset consists of English medical textbooks in plain text (.txt) format.[43] These documents serve as a source of unstructured medical knowledge.

### C. Knowledge Graph Construction

The medical knowledge graph is constructed using the Unified Medical Language System (UMLS) dataset and implemented using the Neo4j graph database (Figure 4). The objective is to model structured medical knowledge through concept nodes and semantically meaningful relationships that reflect clinical reasoning relevant to the USMLE.[20]

#### 1) Graph Database Design

##### a) Node Types

The graph consists of various medical concepts represented as nodes:

- **Anatomy:** Represents body parts and structures.
- **ClinicalScenario:** Captures clinical findings and scenarios.
- **Concept:** General medical concepts.
- **Definition:** Provides textual definitions.
- **Disease:** Encompasses various medical conditions.
- **Drug:** Includes pharmacologic substances and clinical drugs.
- **Procedure:** Covers diagnostic and therapeutic interventions.
- **SemanticType:** Classifies nodes into semantic categories.
- **Symptom:** Represents clinical signs and symptoms.

##### b) Relationship Types

Key relationships include:

###### Clinical Relationships

- `MAY_BE_TREATED_BY`: Links diseases to treatments.
- `HAS_CAUSATIVE_AGENT`: Connects diseases to causes.
- `ASSOCIATED_WITH`: Captures general associations.

###### Disease Relationships

- DISEASE_HAS_FINDING: Links diseases to clinical findings.
- `MANIFESTATION_OF`: Connects symptoms to diseases.

###### Drug Relationships

- `HAS_MECHANISM_OF_ACTION`: Describes pharmacological effects.
- `CONTRAINDICATED_WITH_DISEASE`: Indicates unsafe drug-disease interactions.

#### 2) Graph Construction

##### a) Tools

The graph is implemented in Neo4j for efficient querying and relationship handling.

##### b) Process

The overall graph construction process is illustrated in Figure 6.

- **Node Creation:** Nodes are created for each unique medical concept.
- **Relationship Creation:** Links are established using `MERGE` to avoid duplicates.
- **Definition Integration:** Definitions from MRDEF are linked to nodes.

### D. Vector Store Construction (Pinecone)

The vector store component enables semantic search over unstructured medical content derived from textbooks (Figure 5). We selected Pinecone as our vector store due to its scalability, low-latency retrieval, and support for real-time approximate nearest neighbor (ANN) search optimized for high-dimensional embeddings.This facilitates dense retrieval of relevant textual evidence based on user queries, complementing the structured reasoning provided by the knowledge graph.

**Fig. 5:**
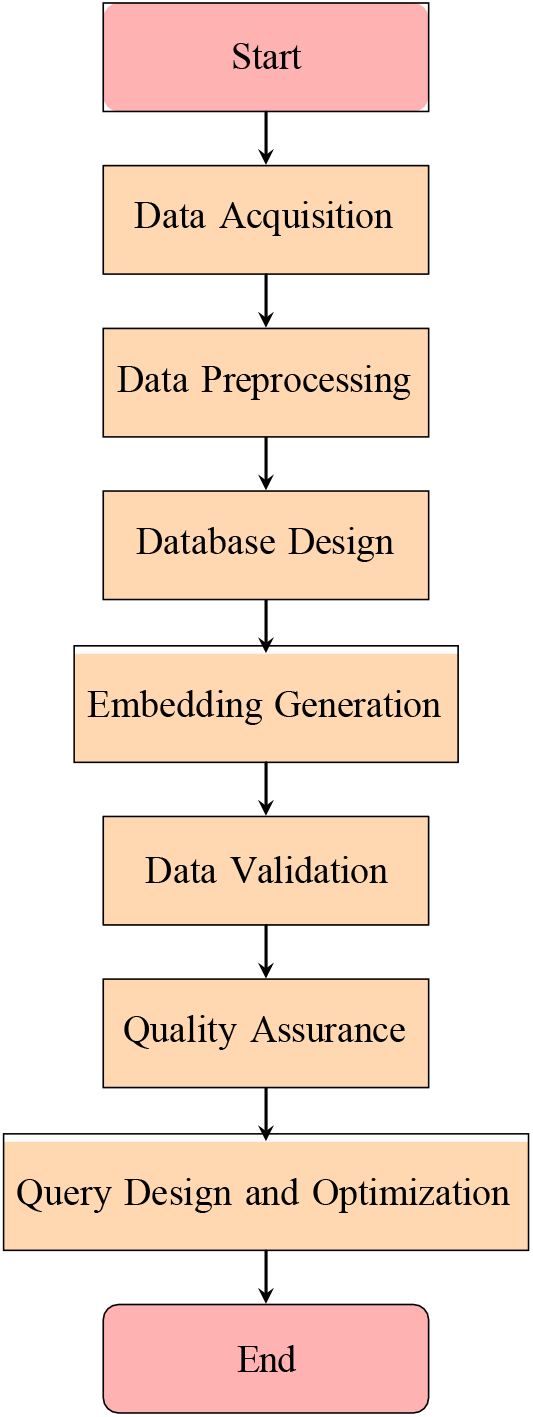
Workflow of Information Retrieval System

**Fig. 6:**
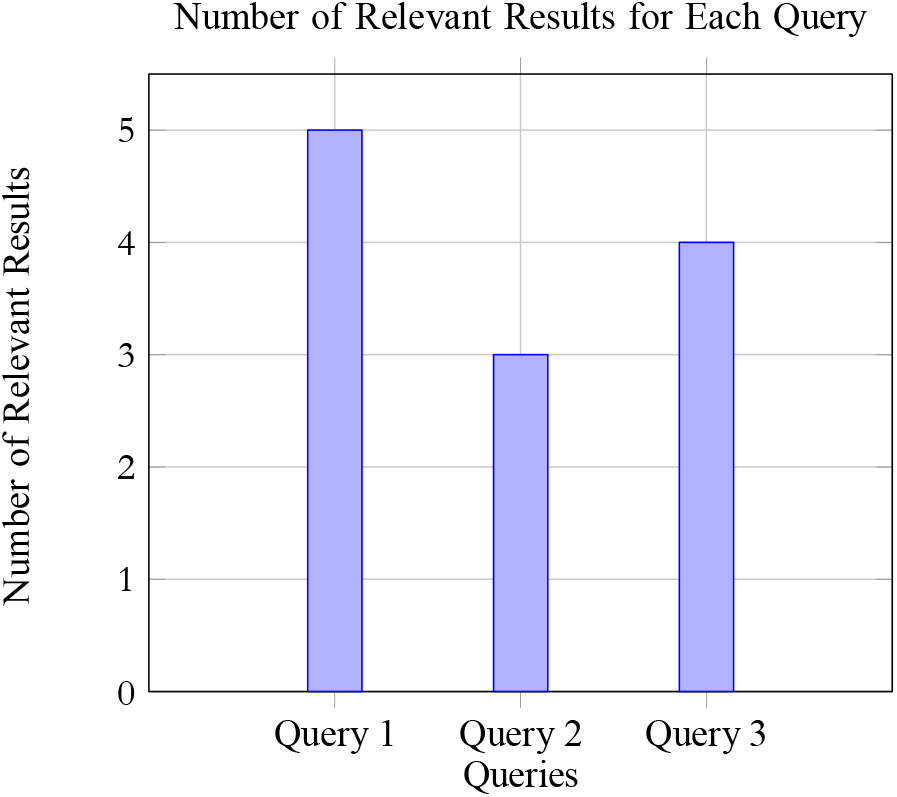
Bar graph showing the number of relevant results for each query.

#### 1) Textbook Dataset and Preprocessing

The input dataset consists of English-language medical textbooks stored in plain text (.txt) format. The following pre-processing steps were implemented:

- **Cleaning:** Non-content elements (e.g., figure/table references) were removed using regular expressions such as Fig.,Table., etc.
- **Sentence Splitting:** Each textbook was split into individual sentences using `RecursiveCharacterTextSplitter` from the LangChain library, configured with a chunk size of 1 and no overlap for maximum granularity.
- **Metadata Generation:** Each sentence was associated with the `file name` as the `source`.

#### 2) Embedding Generation

Semantic embeddings were generated using the `text-embedding-3-large`[41] model from OpenAI. We selected OpenAI’s `gpt-embedding-3-large` model for encoding MedQA textbook content due to its superior performance on semantic similarity benchmarks and broad generalization capabilities across diverse domains, including biomedical text. To optimize the process:

- **Batching:** Sentences were processed in batches of 1000 using the `batch_size` argument to reduce memory overhead.
- **Parallelization:** Embedding generation was distributed using Python’s `concurrent.futures.ThreadPoolExecutor`, allowing multiple batches to be encoded in parallel.
- **Mapping:** Each sentence’s embedding was stored alongside its metadata for downstream retrieval.

#### 3) Vector Store Indexing

The embeddings are indexed in Pinecone, a vector database optimized for similarity search, using the `process_batch` function. The Pinecone index is created with a cosine similarity metric and a dimensionality of 3072, matching the `text-embedding-3-large` embeddings. Each batch of embeddings is upserted into the index with associated metadata, allowing for efficient retrieval based on semantic similarity.

While this pipeline was designed for integration with Pinecone, it is also compatible with other vector stores such as ChromaDB, enabling flexible back-end storage options.

#### 4) Query-Time Embedding and Retrieval

At runtime, user queries are processed using the same embedding model (Figure 6). The query vector is compared against stored sentence vectors using cosine similarity, and the top-*k* results are returned [21].

- **Embedding Consistency:** Ensured via model reuse and fixed preprocessor settings.
- **Top-***k* **Retrieval:** The most relevant sentences are selected for fusion with graph-based context.
- **Relevance Filtering:** Retrieved outputs are post-processed to ensure domain alignment and context completeness.

#### 5) Query Performance

The information retrieval system was tested with a set of predefined queries, including:

- “What are the symptoms of a heart attack?”
- “How does DNA replication work?”
- “What is the role of the immune system?”

For each query, the system successfully retrieved relevant sentences from the database, demonstrating its capability to provide accurate and contextually appropriate responses. The results for each query were as follows:

- Query 1: The system returned 5 relevant sentences, each providing insights into the symptoms of a heart attack. The metadata for these results included the source text-book and a brief summary of each section, enhancing the interpretability of the information presented.
- Query 2: For the query regarding DNA replication, the system retrieved 3 relevant sentences. The embeddings generated for these sentences effectively captured the key concepts related to the process of DNA replication, allowing users to gain a clear understanding of the topic.
- Query 3: The query about the immune system yielded 4 relevant results. The retrieved sentences highlighted various aspects of the immune system’s function, supported by the corresponding metadata that provided context for each piece of information.

## IV. Hybrid Retrieval and Context Fusion Strategy

To answer USMLE-style questions effectively, our system adopts a hybrid retrieval strategy that integrates symbolic reasoning from a knowledge graph with dense retrieval from a vector store. This section describes how relevant information is extracted, ranked, and fused before being passed to the large language model (LLM).

### 1) Extraction of Key Medical Terms from User Query

To initiate the retrieval process from the knowledge graph, the system employs a large language model (LLM) to extract key medical terms from the user’s query. The LLM comprehensively identifies and categorizes terms into predefined medical types such as

- Disease (e.g., diabetes)
- Drug (e.g., penicillin)
- Symptom (e.g., fever)
- Anatomy (e.g., heart)
- Procedure (e.g., ECG)
- Clinical Scenario (e.g., “45-year-old man”)
- Laboratory Value (e.g., “hemoglobin 9 g/dL”)

This categorization facilitates targeted querying of the knowledge graph. The extraction process captures all relevant medical concepts ensuring a robust foundation for graph-based retrieval[4]. The extracted terms for a sample question (Figure 7) will be:

**Fig. 7:**
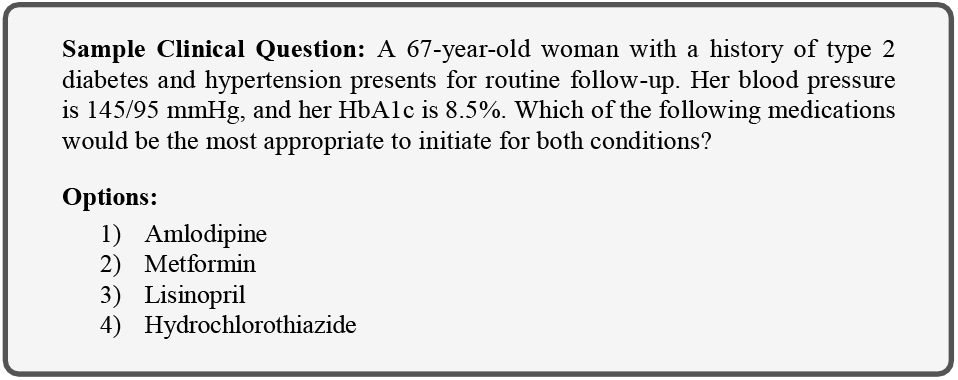
Sample Question for term extraction

- **ClinicalScenario:** 67-year-old woman, routine follow-up
- **Disease:** type 2 diabetes, hypertension
- **LaboratoryValue:** blood pressure 145/95 mmHg, HbA1c 8.5%
- **Drug:** Amlodipine, Metformin, Lisinopril, Hydrochlorothiazide

### 2) Retrieving Concepts

Following term extraction, the system retrieves corresponding concepts from the knowledge graph using a dual-strategy approach:

- **Direct matching:** In direct matching, the system queries the graph for concepts whose terms exactly match or contain the extracted terms, employing case-insensitive comparisons.
- **Vector similarity search:** For terms lacking exact matches, particularly synonyms or semantically related concepts, a vector similarity search is conducted. This method computes embeddings for both the extracted terms and graph concepts using a pre-trained model (e.g., Bio_ClinicalBERT), identifying semantically similar concepts based on cosine similarity.

This combined approach ensures comprehensive concept coverage, bridging textual and semantic gaps in the knowledge graph (Figure 8).[22]

**Fig. 8:**
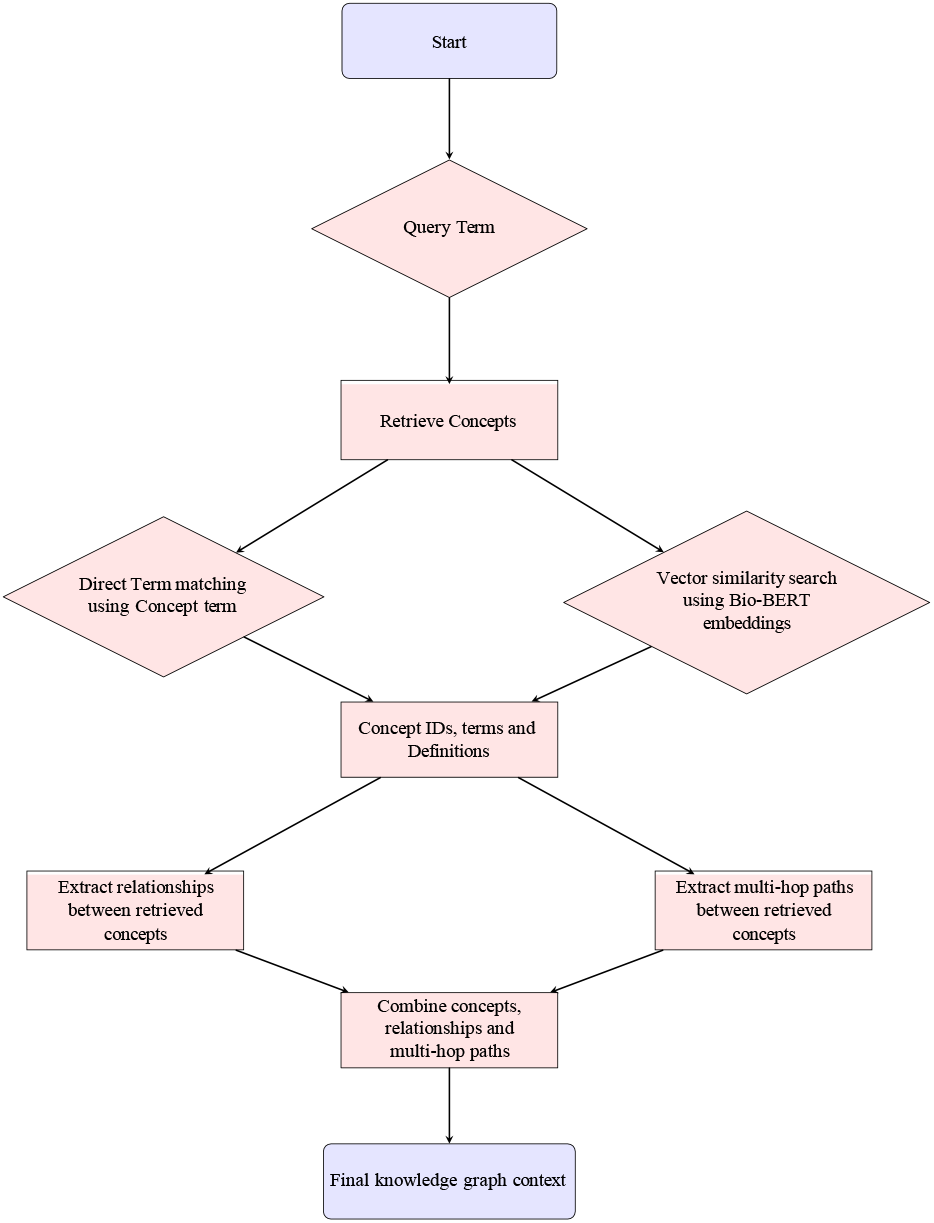
Pipeline for retrieving concepts, relationships and multi-hop paths from knowledge graph

### 3) Retrieving Relationships

The system retrieves all relationships connecting these concepts within the knowledge graph. It queries for relationships where either the source or target concept matches the retrieved concepts’ unique identifiers (CUIs), without filtering by relationship type.

This inclusive strategy captures a broad spectrum of connections, including causal links (e.g., “CAUSE_OF”), treatment associations (e.g., “MAY_TREAT”), and symptomatic relationships (e.g., “DISEASE_HAS_FINDING”). By preserving all relationship types, the system ensures a rich contextual dataset, avoiding the loss of potentially pertinent medical insights, which is critical for answering complex clinical queries.[23]

### 4) Retrieval of Multi-Hop Paths

The system also searches for multi-hop paths between pairs of retrieved concepts, up to a configurable maximum depth (e.g., 3 hops). This process employs a graph traversal algorithm that identifies paths connecting concepts via any relationship type, without restrictions, to maximize discovery of meaningful associations. [24]

For example, a path might link a symptom to a disease through an intermediate physiological process, such as “(chest pain)-[DISEASE_HAS_FINDING] → (myocardial infarction)-[CAUSE_OF] → (coronary artery occlusion).” These multi-hop paths provide additional context and insight, enhancing the system’s ability to address intricate medical scenarios beyond direct relationships.

### 5) Re-Ranking Strategy

Given the large volume of data retrieved from the knowledge graph, a re-ranking strategy is employed to prioritize the most relevant concepts, relationships, and multi-hop paths based on their alignment with the user’s query. This ensures that the system focuses on the most pertinent information, enhancing the accuracy and contextual relevance of the generated answers.

The relevance score for a concept *c* is calculated as a weighted combination of its original confidence score and its semantic similarity to the query:

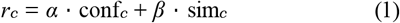

where:

- conf_*c*_: The original confidence score of the concept (e.g., from initial retrieval or vector similarity).
- sim_*c*_: The semantic similarity between the concept and the query, computed using cosine similarity of their embeddings.
- *α* = 0.3, *β* = 0.7: Weights that prioritize semantic relevance while considering the initial confidence.

This approach ensures that concepts are ranked based on both their initial retrieval strength and their contextual alignment with the query.

For relationships, the relevance score *r*_rel_ is determined by a weighted sum of the semantic similarity of the relationship statement and the relevance scores of its source and target concepts:

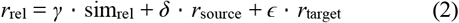

where:

- sim_rel_: The semantic similarity between the relationship statement (e.g., “A relates to B”) and the query.
- *r*_source_, *r*_target_: The relevance scores of the source and target concepts, computed using the concept re-ranking formula.
- *γ* = 0.4, *δ* = 0.3, *ϵ* = 0.3: Weights that emphasize the relationship’s relevance while considering the importance of connected concepts.

This formula promotes relationships that are semantically aligned with the query and connect highly relevant concepts. For multi-hop paths, the relevance score *r*_*p*_ combines semantic similarity with a penalty for path length to favor concise yet informative connections:

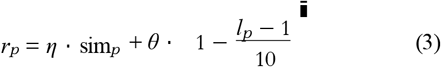

where:

- sim_*p*_: The semantic similarity between the path’s description and the query.
- *l*_*p*_: The length of the path (number of hops).
- *η* = 0.8, *θ* = 0.2: Weights that prioritize semantic relevance while applying a small penalty for longer paths.
- 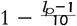: A length penalty term that decreases as path length increases.

This ensures that multi-hop paths are both relevant and concise, enhancing their utility in providing meaningful connections.

By applying these re-ranking strategies, the system effectively prioritizes the most relevant information from the knowledge graph, ensuring that the generated answers are both accurate and contextually appropriate. The use of semantic similarity and weighted scoring allows for a nuanced selection of concepts, relationships, and paths, ultimately improving the system’s ability to address complex clinical queries.[25]

#### A. Vector-Based Retrieval and Re-Ranking

This subsection outlines the process of retrieving and reranking relevant medical text passages from a Pinecone vector store using vector-based methods. The approach involves generating vector embeddings for the query, querying the Pinecone index to retrieve similar text chunks, combining similar chunks to form coherent passages, which is later given as context to the LLM to generate the final response (Figure 9).

**Fig. 9:**
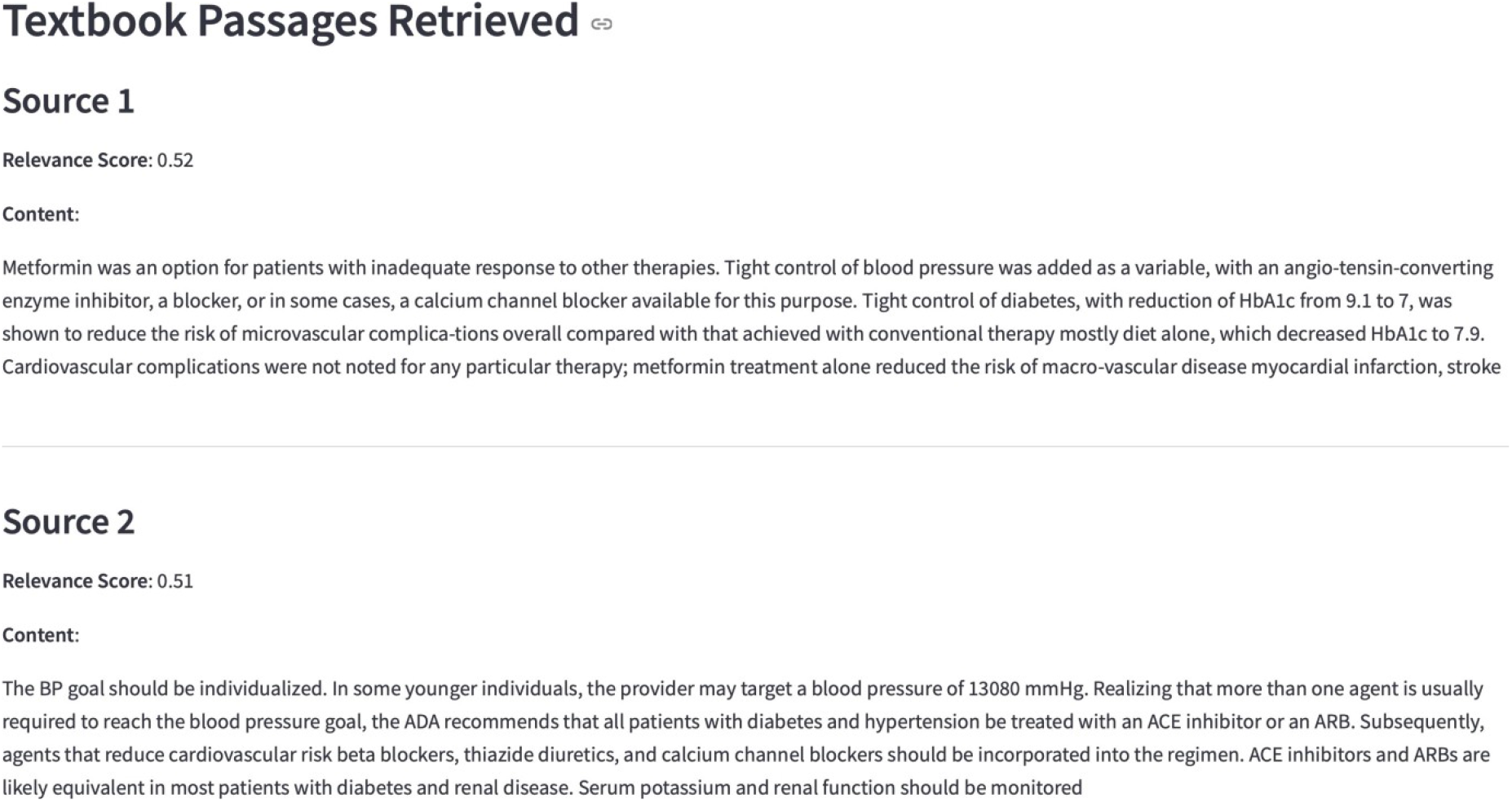
Retrieved relevant textbook passages from Pinecone for sample question

### 1) Vector Embedding Generation

The user’s query is transformed into a vector embedding using OpenAI’s `gpt-embedding-3-large` model. This embedding, denoted as **e**_*q*_, captures the semantic meaning of the query in a high-dimensional vector space. The embedding function can be represented as:

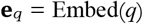

where *q* is the query text, and Embed is the embedding model provided by OpenAI.

### 2) Pinecone Index Querying

The query embedding **e**_*q*_ is used to search the Pinecone index, named `medical-textbook-embeddings`, for the top-*k* most similar vectors (set to 8 in the code). Similarity is measured using cosine similarity, defined as:

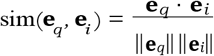

where **e**_*i*_ is the embedding of the *i*-th text chunk stored in the index. Retrieved chunks are filtered to include only those with a similarity score above a threshold of 0.5, ensuring relevance to the query.

### 3) Similarity-Based Chunk Combination

To enhance coherence, similar text chunks are combined based on their textual similarity. The similarity between two chunks is calculated using the SequenceMatcher ratio from Python’s `difflib` module:

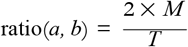

where *M* is the number of matching characters, and *T* is the total number of characters in both texts *a* and *b*. Chunks are grouped if their similarity ratio exceeds a threshold of 0.3 or if there is direct textual overlap (e.g., one chunk’s text appears within another). This process consolidates related information into coherent passages, preserving contextual integrity.[26]

#### A. Context-Fusion and Response Generation

In this section, we detail the process of fusing context from both the knowledge graph and vector store and generating a coherent, evidence-based response using a large language model (LLM).We make use of prompt-based response synthesis with GPT-4o models to ensure that answers are strictly grounded in the retrieved medical references. [27][28]

### 1) Context Formatting

To prepare the retrieved context for LLM processing, data from the knowledge graph and vector store is formatted into a structured, readable format. Knowledge graph information, including concepts, relationships, and multi-hop paths, is arranged into clear sections with identifiers and descriptions. Vector store textbook passages are formatted with source tags, relevance scores, and combined into coherent chunks based on similarity. This fused context provides a comprehensive, readable input for generating evidence-based responses.

### 2) Response Synthesis using LLM

For response generation, we opted for GPT-4o-mini[42] as a it provides strong reasoning capabilities while being significantly more efficient than larger LLMs, allowing us to evaluate the impact of context quality independently of model scale. Answers are synthesized under three strategies, each guided by distinct prompts:

- **LLM-Only Response**: This approach relies solely on the LLM’s internal knowledge, without external context. The prompt instructs the LLM to provide a detailed answer with reasoning, differential analysis, and clinical context, selecting the best multiple-choice option if provided.
- **Context-Strict Response**: This strategy restricts the LLM to using only the provided knowledge graph and textbook evidence, prohibiting external knowledge. The prompt, depicted in Figure 10, enforces a structured response format including reasoning, evidence integration, and clinical context, prioritizing graph data over textbook passages. Citations to specific concept IDs and exact textbook quotes are mandatory.
- **LLM-Informed Response**: This approach integrates the LLM’s expertise with the fused context, prioritizing graph evidence, followed by textbook data, and using internal knowledge only when gaps remain. This prompt mirrors the context-strict format but permits expert reasoning to resolve ambiguities, enhancing answer robustness.

**Fig. 10:**
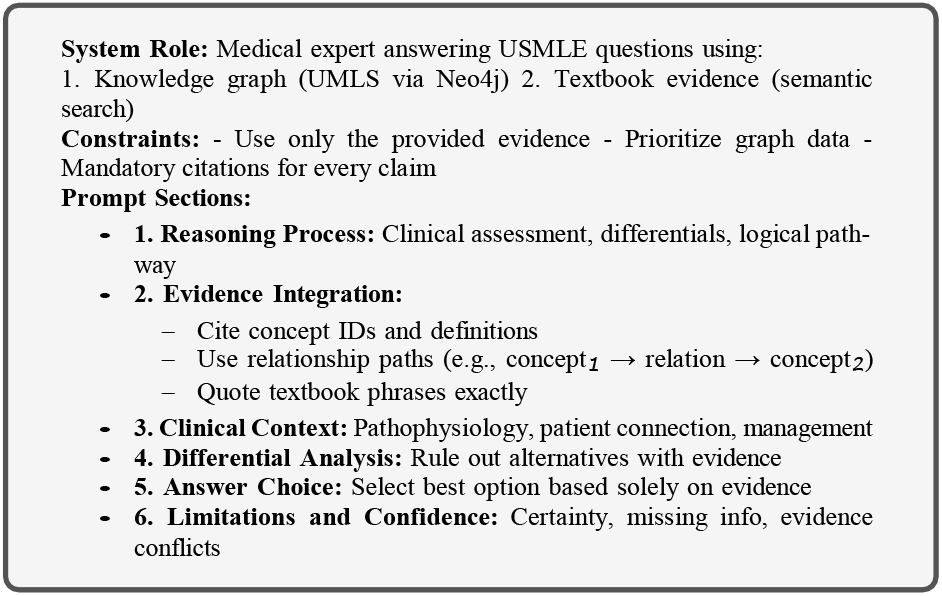
Structured overview of the prompt used for context-strict LLM responses.

The rationale behind adopting three response strategies is to enable comparative evaluation of answer quality, factual accuracy, and citation relevance across varying levels of contextual augmentation. Specifically, we aim to observe how the presence or absence of knowledge graph and textbook evidence affects the LLM’s clinical reasoning and citation behavior. The answer generated for the sample question(Figure 9) by the Context-Strict prompt is shown in Figure 11.

**Fig. 11:**
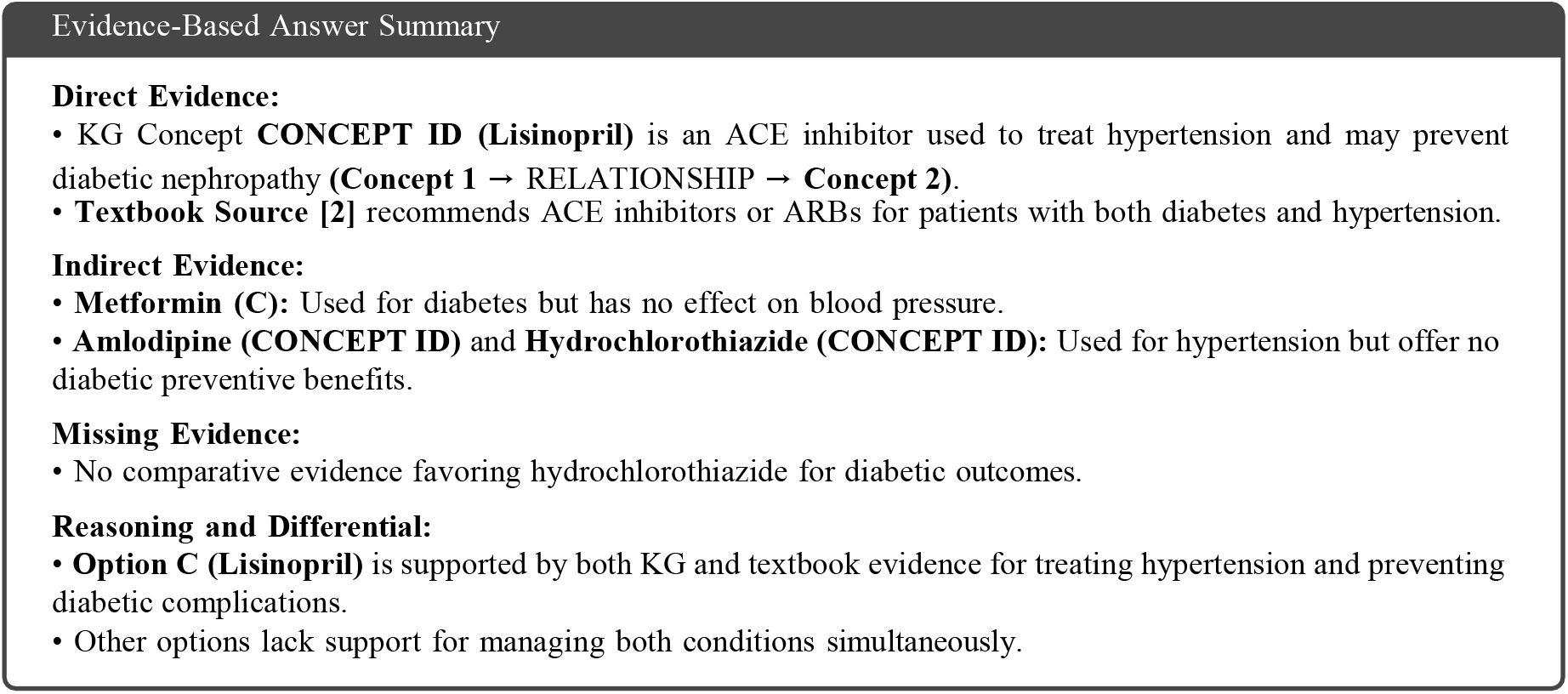
Sample Context-Strict Answer

## V. Ablation Study

An ablation study was carried out using 30 randomly selected questions from the USQuestionBank JSON file provided by the MedQA dataset [43]. The primary goal of this experiment was to establish a proof-of-concept for our proposed pipeline. Specifically, the study evaluates and compares the performance of the following three strategies:

- LLM Only
- Context Strict
- LLM Informed

### A. Evaluation Metrics and Technique

In the absence of structured ground truth for evaluating the reasoning components of USMLE-style medical questions, we adopted an approach where Large Language Models (LLMs) assess the responses. This method draws inspiration from the LLM-Guided Evaluation framework discussed by Arthur AI [40], which explores using LLMs to evaluate the outputs of other LLMs, highlighting both the potential and challenges of this approach.

The GraphRAG system employs a comprehensive evaluation methodology that integrates multiple assessment approaches to measure the quality, correctness, and evidence-based reasoning capabilities of the system. The evaluation framework comprises several interconnected components:

#### 1) Evidence-Based Assessment

The evaluation prioritizes both the evidence quality and factual correctness of generated answers:

##### Knowledge Graph Coverage Analysis

- **Concept Coverage (%):** Measures the percentage of key medical terms from the question present in the knowledge graph.
- **Relationship Utilization:** Evaluates the effectiveness of leveraging connections between medical concepts.
- **– Multihop Path Relevance:** Assesses the relevance and quality of indirect connections discovered through graph traversal.

##### Context Contribution Metrics

Metrics to quantify the contribution of retrieved context include:

- **Value Added Score (0–10):** Quantifies the improvement in answer quality due to retrieved context compared to LLM-only answers.
- **Citation Analysis:** Evaluates how effectively citations from the knowledge graph are incorporated into the answers.
- **Hallucination Detection:** Identifies and quantifies unsupported claims in the answers.

A combined evidence score ranging from 0 to 10 is assigned based on the overall quality of the aforementioned evidence-related parameters present in the final response.

#### 2) Correctness Score (0–10)

Evaluates the factual accuracy of the generated answers.

- Compares answers against reference solutions for USMLE-style questions.
- Uses semantic similarity matching for nuanced assessment.
- Considers partial matches and conceptual accuracy.

#### 3) Combined Score (0-10)

The combined score is computed as a weighted average of the evidence and correctness scores according to the following equation:

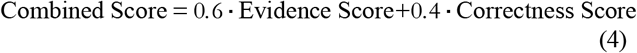

These weights were chosen to emphasize evidence-based reasoning, reflecting the critical importance of verifiable justifications in medical question-answering. Assigning a higher weight (0.6) to the evidence score underscores the system’s primary objective: producing transparent, traceable answers supported by explicit medical knowledge. Meanwhile, a weight of 0.4 is assigned to correctness to ensure that factual accuracy remains an integral component of overall answer quality.

### A. Results and Analysis

As shown in Figure 12, the LLM-Informed method consistently outperforms both the LLM-Only and Context-Strict approaches, achieving a 15 percent improvement in overall accuracy compared to the LLM-Only baseline. This confirms that the integration of structured knowledge with LLM capabilities produces superior results.

**Fig. 12:**
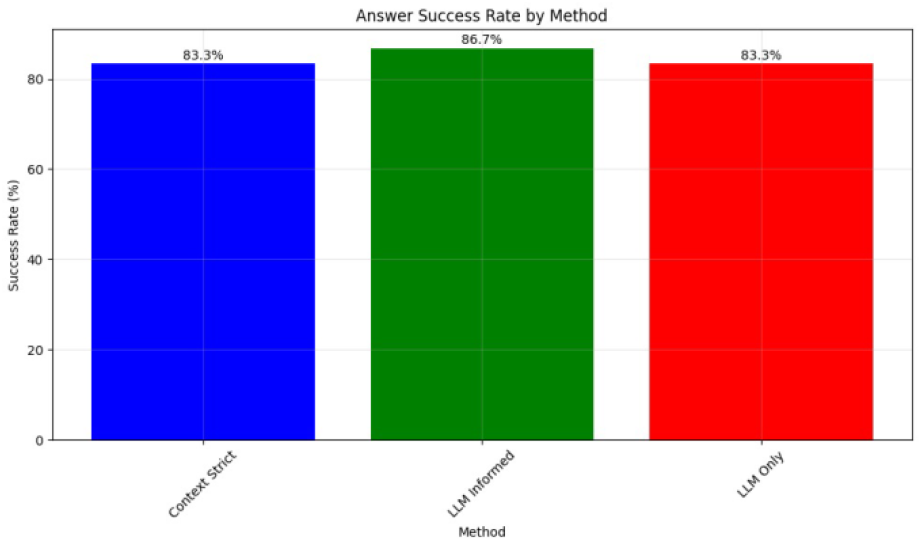
Success rates across different answering methods

The comparative analysis (Figure 13) reveals that the LLM Informed method achieved the highest overall performance, excelling in both correctness and evidence-based scoring. Context Strict also performed well but slightly lagged in evidence alignment. In contrast, the LLM Only method, despite high correctness, showed poor evidence scores, resulting in the lowest total performance.

**Fig. 13:**
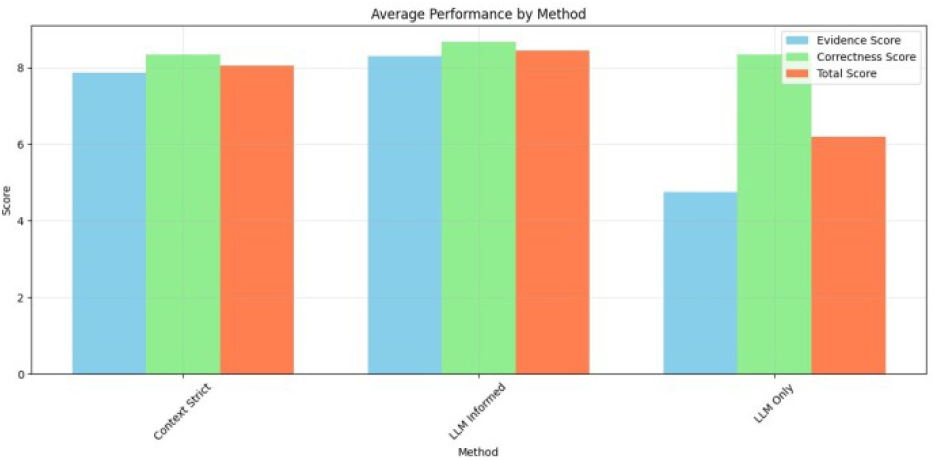
Comparison between evidence quality and correctness scores across different methods.

The boxplot analysis in Figure 14 demonstrates that the LLM-Informed strategy consistently outperforms others, achieving the highest median score with minimal variability, indicating both accuracy and reliability. The Context-Strict approach, while often effective, shows greater variability and occasional failure when evidence is sparse. In contrast, the LLM-Only method performs consistently lower, with limited adaptability across questions. These results highlight the critical role of contextual evidence in enhancing both the quality and consistency of medical question answering.

**Fig. 14:**
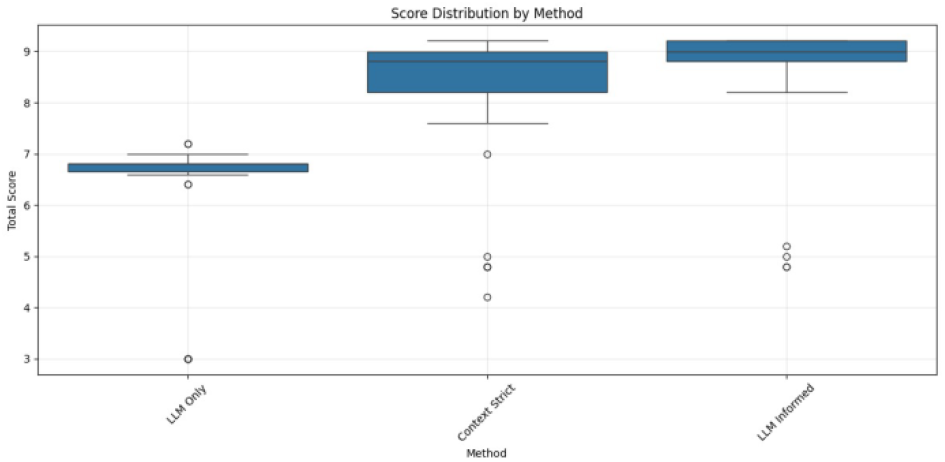
Distribution of combined scores across the three answering methods.

Overall, the Context-Strict and LLM-Informed strategies outperform the LLM-Only approach, particularly in citation quality and, in few cases, factual correctness (refer to GitHub: https://github.com/Tharun2331/GRAPHRAG/tree/master/screenshots for detailed results). Our findings show that integrating structured and unstructured evidence can enhance the performance of smaller LLMs, even when using a relatively limited external knowledge base. As the external database expands, we anticipate the potential to further improve performance, including for larger models, by providing more comprehensive and relevant context.

## VI. Conclusion and Future Work

One of the core contributions of our work lies in the integration of structured (Neo4j/UMLS) and unstructured (text-book) knowledge to improve the transparency and traceability of LLM-generated answers in the medical domain. While general-purpose language models are capable of generating plausible-sounding citations, these are often not grounded in real-time retrieval and may include hallucinated references. In contrast, our system enforces evidence grounding through explicit retrieval, re-ranking, and source attribution. This guarantees that every cited concept ID or textbook phrase can be traced back to a verified source in the underlying knowledge graph or vector store. Such traceability is especially important for high-stakes domains like clinical decision-making, where factual correctness alone is insufficient without supporting justification.

Empirically, we find that our pipeline consistently outperforms smaller LLM baselines such as GPT-4o-mini, particularly in generating explainable, step-by-step responses with verifiable citations. However, our performance does not yet surpass that of larger models like GPT-4.5, especially in openended reasoning and nuanced clinical judgment. We attribute this to the limited scale of our retrieval corpus and the current simplicity of our ranking heuristics. Nevertheless, our system presents a scalable framework for integrating verifiable knowledge into LLM pipelines, and we believe that with expanded datasets, improved graph traversal, and learning-to-rank methods, it can provide a competitive alternative to relying purely on larger pretrained models.

## Data Availability

All data and programming of this project is available at the public github

https://github.com/Tharun2331/GRAPHRAG

## Acknowledgment

The first, second, and third authors would like to acknowledge the support provided by the fourth author, Dr. Sabah Mohammed of Department of Computer Science, Lakehead University, for his invaluable guidance throughout this research project. The experimentation and implementation details carried out by the first three authors have been published and are available on Github: https://github.com/Tharun2331/GRAPHRAG.

## APPENDIX

The initial phase of this project involved evaluating a range of transformer-based and traditional models—such as BERT Large, BioBERT, RoBERTa, and SPARQL-based systems—on two prominent medical QA datasets (GBaker/MedQA-USMLE-4-options and Shubham09/medqa). These experiments helped identify performance limitations across models and datasets, especially in terms of generalization and domain adaptation, as indicated by BLEU scores. Additionally, a structured evaluation framework was established using expert-derived scoring rubrics and baseline answer comparisons. This foundational work not only guided the decision to adopt a GraphRAG-based pipeline but also provided critical benchmarks for validating improvements in answer quality through integrated knowledge graph and retrieval-augmented generation techniques. For more detailed implementation, refer to: https://github.com/kushal-ml/MS_Project_Fall24/tree/Tharun.

### APPENDIX A

This appendix summarizes the implementation results of multiple transformer-based and traditional models on two datasets—GBaker/MedQA-USMLE-4-options and Shubham09/medqa—along with references to relevant supporting studies.

#### Model Performance on GBaker/MedQA-USMLE-4-options

**TABLE II:**
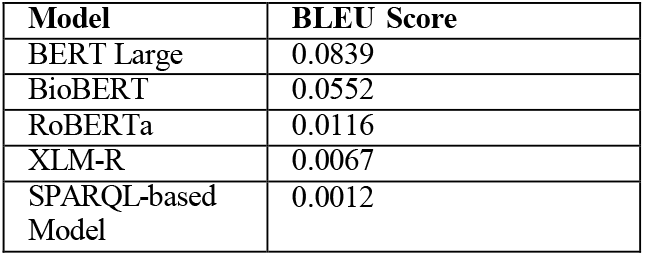
BLEU Scores on GBaker/MedQA-USMLE-4-options.

#### Model Performance on Shubham09/medqa

**TABLE III:**
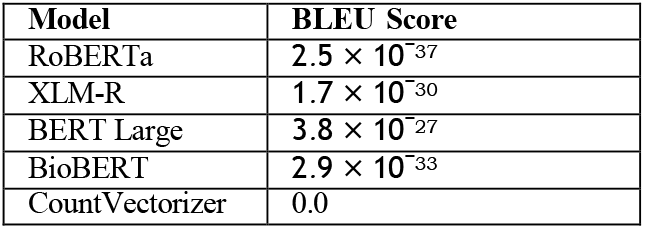
BLEU Scores on Shubham09/medqa.

#### Relevant Studies and Literature

**TABLE IV:**
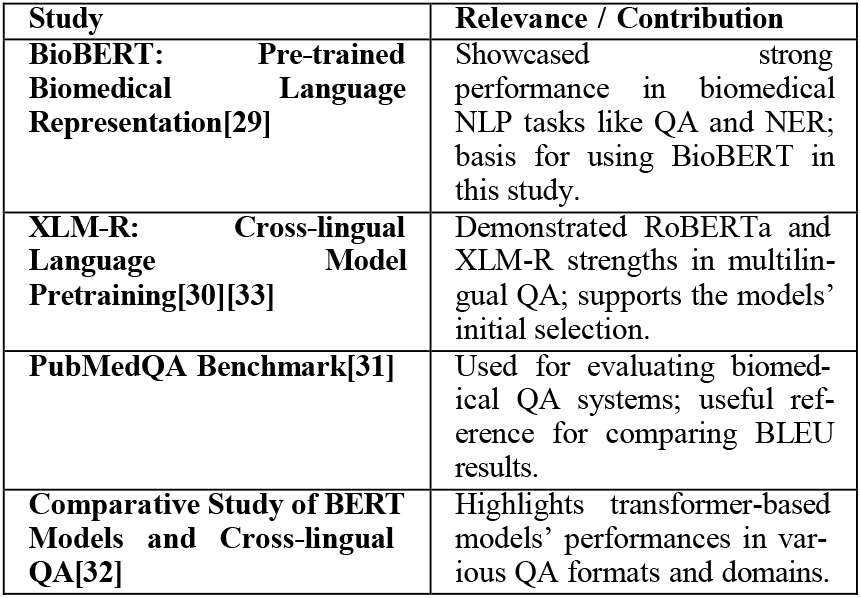
Relevant Studies and Their Contribution to the Project with Access Links.

##### Observations

- **Dataset Sensitivity:** Performance varied significantly by dataset despite similar model architectures.
- **Generalization Gap:** Drop in BLEU scores on Shubham09/medqa highlights limitations in transferability.
- **Domain-Specific Adaptation:** Pretraining or fine-tuning on similar domain data remains essential for better outcomes.

### APPENDIX B

This appendix provides an overview of the evaluation framework used for answer quality assessment, including sample questions, scoring criteria, toolset, and a comparison of generated versus ground truth responses.

**TABLE V:**
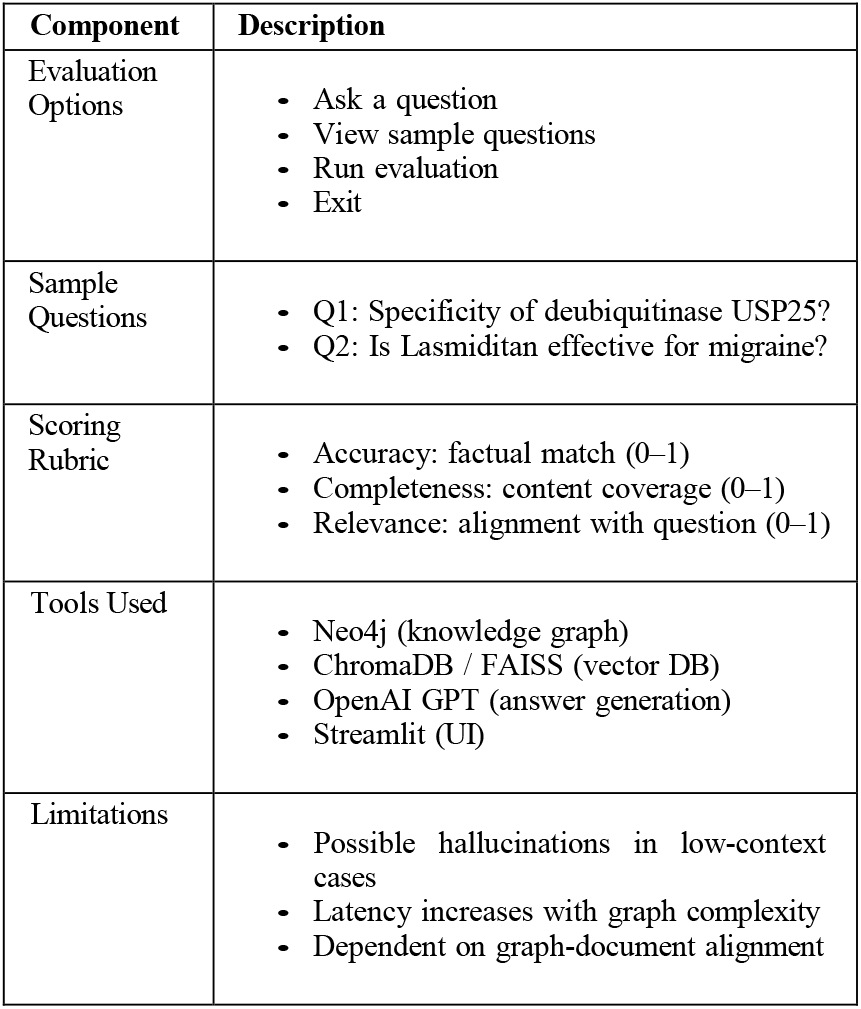
Evaluation Framework Overview.

**TABLE VI:** Generated vs Ground Truth Answers and Scoring Summary.

| Q. No. | Generated Answer (Summary) | Ground Truth Answer (Summary) | Acc. | Comp. | Rel. |
| --- | --- | --- | --- | --- | --- |
| Q1 | USP25 targets K48/K63 chains, regulates immune and DNA repair; specificity due to context | USP25 hydrolyzes K48/K63 chains; involved in immunity, degradation | 0.50 | 0.50 | 0.50 |
| Q2 | Lasmiditan approved for acute migraine; serotonin receptor agonist; effective in trials | Lasmiditan shown effective in migraine trials | 0.75 | 0.75 | 1.00 |
| Avg. | – | – | 0.62 | 0.62 | 0.75 |

**TABLE VII:** Relevant Research Papers, Their Relevance, and Access Links.

| Study | Relevance / Contribution |
| --- | --- |
| **Retrieval-Augmented Generation for Knowledge-Intensive NLP Tasks[34]** | Foundation for combining retrieval and generation in GraphRAG. |
| **Graph Retrieval-Augmented Language Models[35]** | Guides integration of graph-based retrieval for improving answer quality in GraphRAG. |
| **BioBERT: A Pre-trained Biomedical Language Representation Model for Biomedical Text Mining[29]** | Used as a baseline for medical domain-specific answer evaluation. |
| **ROUGE: A Package for Automatic Evaluation of Summaries[36]** | Basis for evaluating answer relevance and overlap. |
| **BLEU: a Method for Automatic Evaluation of Machine Translation[37]** | Supports the use of n-gram overlap to evaluate answer accuracy. |
| **Question Answering over Knowledge Graphs[38]** | Informs the use of knowledge graphs (Neo4j) for structured answer retrieval in medical queries. |
| **Combining Structured and Unstructured Data for Biomedical QA[39]** | Encourages integrating knowledge graphs and text documents for more contextually accurate answers. |

## Notes

### Competing Interest Statement

The authors have declared no competing interest.

### Funding Statement

The Last two authors MITACS Accelerates Grant

### Author Declarations

The source of data are open and available on the project Github: https://github.com/Tharun2331/GRAPHRAG

